# Maternal and neonatal IgG against *Klebsiella pneumoniae* are associated with broad protection from neonatal sepsis: a case-control study of hospitalized neonates in Botswana

**DOI:** 10.1101/2024.05.28.24308042

**Authors:** Siqi Linsey Zhang, Carolyn M. McGann, Tereza Duranova, Jonathan Strysko, Andrew P. Steenhoff, Alemayehu Gezmu, Britt Nakstad, Tonya Arscott-Mills, One Bayani, Banno Moorad, Nametso Tlhako, Melissa Richard-Greenblatt, Paul J. Planet, Susan E. Coffin, Michael A. Silverman

**Author notes:** These authors contributed equally to this paper (Siqi Linsey Zhang, Carolyn M. McGann).

## Abstract

Sepsis is the leading postnatal cause of neonatal mortality worldwide. Globally *Klebsiella pneumoniae* is the leading cause of sepsis in hospitalized neonates. This study reports development and evaluation of ELISA for anti-*Klebsiella* IgG using dried blood spot samples and evaluates the association of anti-*Klebsiella* IgG (anti-Kleb IgG) antibodies in maternal and neonatal samples and the risk of neonatal sepsis. Neonates and their mothers were enrolled at 0-96 hours of life in the neonatal unit of a tertiary referral hospital in Gaborone, Botswana and followed until death or discharge to assess for episodes of blood culture-confirmed neonatal sepsis. Neonates with sepsis had significantly lower levels of *Kleb-*IgG compared to neonates who did not develop sepsis (Mann-Whitney U, p=0.012). Similarly, samples from mothers of neonates who developed sepsis tended to have less *Kleb-*IgG compared to mothers of controls (p=0.06). The inverse correlation between *Kleb-IgG* levels and all-cause bacteremia suggests that maternal *Kleb-* IgG is broadly protective through cross-reactivity with common bacterial epitopes. These data support the continued use of immunoglobulin assays using DBS samples to explore the role of passive immunity on neonatal sepsis risk and reaffirm the critical need for research supporting the development of maternal vaccines for neonatal sepsis.

## INTRODUCTION

Sepsis is the leading postnatal cause of neonatal deaths, accounting for nearly one million deaths annually.(1) The incidence, causative pathogens and mortality attributable to neonatal sepsis vary globally.(2) In high-income countries, the most common causes of early-onset sepsis (EOS) are Group B streptococcus (GBS) and *Escherichia coli* whereas the vast majority of late-onset sepsis (LOS) cases are caused by Gram-positive organisms such as coagulase-negative staphylococci (CoNS) and *Staphylococcus aureus*.(3–5) In contrast, both EOS and LOS are predominantly caused by Gram-negative organisms in low- and middle-income countries (LMIC).(6–8) In Botswana and other sub-Saharan African countries, Gram-negative sepsis accounts for about 60% of neonatal sepsis, with a predominance of *Klebsiella* spp.(9–11) Notably, over three-quarters of the global neonatal sepsis deaths occur in LMIC signifying the critical unmet need to better understand the immune system features that predispose to, or protect against neonatal sepsis in these settings.(12) There has been a global focus on strategies to prevent neonatal infection, particularly GBS, including prophylactic antibiotics and a novel vaccine; however, geographic variation in the causes of neonatal infections highlights the need to better understand the risk factors for Gram-negative sepsis in neonates.

Since neonatal innate and adaptive immune systems are immature at birth, the immunoglobulins obtained through transplacental transfer and breastmilk provide critical protection from sepsis.(13–16) Immunoglobulin G (IgG) is the primary antibody transplacentally transferred and plays an essential role in protecting neonates from infection.(17, 18) Although the transfer of IgG begins at 13 weeks of gestation, it accelerates significantly after 36 weeks. Thus preterm, as compared to term, neonates are less protected by passive immunity contributing to their higher risk of sepsis.(18, 19) We hypothesized that lack of maternal IgG antibodies that target specific pathogenic bacteria may predispose neonates to develop sepsis. We developed an enzyme-linked immunosorbent assay (ELISA) to evaluate the degree to which the risk of neonatal sepsis is associated with the level of IgG from maternal and neonatal serum capable of binding *K. pneumoniae* (*Kleb-*IgG).

## METHODS

### Setting and population

This study used samples collected from a cohort of 467 neonates admitted to the neonatal unit at a tertiary referral hospital in Gaborone, Botswana from November 1, 2020 – December 31, 2021. Participants were eligible for enrollment in the parent study if admitted to the neonatal unit within 3 days of life and were followed until discharge or death. A research nurse approached mothers after delivery to obtain written informed consent in Setswana or English. The study was approved by the institutional review boards of the Botswana Ministry of Health Research and Development Committee (protocol 13/18/1, Jan. 16, 2020), the University of Botswana (protocol 147, Dec. 13, 2019), the University of Pennsylvania (protocol 833786, April 22, 2020), the hospital IRB (protocol 708, April 2, 2020) and Children’s Hospital of Philadelphia (protocol 19-016848, July 25, 2020). Cases of sepsis were matched with controls also admitted to the neonatal unit but without sepsis in a 1:4 ratio based on nearest sample collection date and gestational age. Subjects unable to be matched by GA were matched with the nearest GA. Subjects with missing data or samples were excluded. Patients and the public were not included in the design or implementation of this study.

### Sample collection

Dried blood spots (DBS) were collected by heel stick from enrolled neonates and finger stick from mothers timed with a clinically indicated blood sample and stored on filter paper (Whatman 903, Cytiva, USA). We were unable to stick a second time if a patient stopped bleeding, thus collected volumes varied. Blood spots were dried at room temperature for three hours and placed in plastic bags with desiccant packs. They were stored at room temperature before shipment (13-26°C) and at ™20°C after shipment from Botswana to the United States to enhance stability of the samples. Samples were transported at room temperature in two shipments: the first was shipped 6-8 months after collection date and the second shipment contained both neonatal and maternal DBS collected within 4 months. Sample integrity was established prior to analysis.

Blood cultures were drawn at the discretion of the clinical team for suspected clinical sepsis using conventional clinical criteria.(20) Samples were incubated on an automated blood culture system (BACT/ALERT, bioMerieux, Marcy l’Etoile, France) for up to 5 days or until blood culture positivity. Neonatal sepsis was defined as a positive blood culture. CoNS spp. and other skin commensals were deemed contaminants unless ≥2 cultures within 7 days grew this organism.(21)

### Extraction of antibodies from dried blood spots

To extract antibodies from DBS, a 6.35-mm-diameter punch of a saturated blood spot was placed in an Eppendorf tube with 500 μl of elution buffer (phosphate buffered saline [PBS] with 0.1% BSA and protease inhibitor cocktail, Roche, catalog no. 04693159001) then were incubated overnight on a shaker at 4°C. Samples were then centrifuged for 5 mins at 8,000g at 4°C to pellet debris and the supernatants were collected and stored at ™20°C.

### Immunoglobulin quantification for DBS eluates

To quantify the total amount of IgG and IgA present in each eluate, ELISA was performed using Bethyl Human ELISA Quantitation Set (E80-100, E80-102, E80-104) per manufacturer instructions (Bethyl Laboratories, Montgomery, TX, USA).

### Bacterial-binding IgG quantification for DBS eluates using ELISA

We developed an ELISA using bacteria-coated plates to determine the degree to which neonatal antibodies bind to bacterial surface epitopes of *Klebsiella pneumoniae* isolate MF391. This isolate was obtained from a blood culture sample in Botswana. The isolate was subcultured to a Luria broth (LB) solid media and incubated at 37°C overnight. A single colony was selected and transferred to 5 mL of LB to establish a liquid culture. The following day, the culture was centrifuged (10-minute at 4,000xg), fixed with 4% paraformaldehyde for 20 minutes and washed three times in PBS (pH 7.4). The pellet was resuspended in PBS and diluted to achieve an optical density of 1.0. A 100 μl volume of bacteria was added to each well of a 96-well ELISA plate (Thermo Scientific Nunc, 442404). The plate was incubated overnight at 4°C then washed (0.07 M NaCl and 0.025% Tween 20 dissolved in Tris buffer) and blocked (0.007 M NaCl and 1% BSA in Tris) for 30 minutes at room temperature. DBS eluates were diluted (0.007 M NaCl, 1% BSA, 0.025% Tween 20 in Tris buffer) to a uniform IgG concentration of 0.4 ng/μl. 100 μl of the diluted DBS samples were added to each well and incubated overnight at 4°C. The wells were then washed and incubated with horseradish peroxidase-conjugated anti-IgG antibody (1 mg/ml diluted 1:150,000) for an hour and developed for 15 minutes in 3,3’,5,5’-tetramethylbenzidine (TMB) with 0.18M H_2_SO_4_ as the stopping reagent. The absorbance at 450nm was measured immediately on an Enspire Multimode Plate Reader.

A standard unit (SU) was established using reference serum from a healthy adult to establish a standard curve to normalize all samples. A standard ELISA was used to determine the IgG concentration of this serum sample. The sample was then serially diluted to create a standard curve for each experiment. We defined one SU as the optical density of 0.5 μg/mL IgG of the reference serum. Due to variability in the amount of blood collected on each card, we normalized the microbial ELISA assay by adding the same amount of IgG (i.e., 50 ng, an amount determined by titration experiments) to each well to determine the relative amount of *Klebsiella*-binding IgG antibodies from each sample.

To determine the amount of IgG binding to common bacterial epitopes, we used *E. coli* lipopolysaccharide and *Salmonella typhimurium* flagellin. A 100 μl volume of diluted flagellin or LPS (1 μg/mL) was added to each well of a 96-well ELISA plate and incubated overnight at 4°C. The remainder of the ELISA was performed as described above. The secondary anti-LPS and anti-flagellin antibodies were diluted 1:5000.

### Statistical analysis

Nonparametric Mann-Whitney U tests were used to compare *Kleb-*IgG levels and demographic characteristics between neonates with sepsis and healthy controls. A Kruskal-Wallis test was used to compare gestational age categories between the two groups. Simple linear regression was used to evaluate trends in antibody levels. We conducted an exploratory analysis using logistic regression to evaluate the association of *Kleb-*IgG levels with sepsis while controlling for both gestational age and days from storage to sample analysis. We conducted an additional exploratory analysis using simple linear regression to explore the association between maternal and neonatal antibody levels among neonates with and without sepsis to evaluate whether antibody transfer differed. Results were considered significant using a two-sided p value (p<0.05).

## RESULTS

### Study participant characteristics

Of the 467 patients enrolled, 30 (6%) experienced bloodstream infections, 116 (24%) had neonatal DBS shipped recently and available for analysis of whom 8 were diagnosed with sepsis (**Fig. 1**). The most common organisms cultured from the neonates with sepsis were *Acinetobacter* spp. and *Klebsiella* spp. which was similar to the etiologies of sepsis in the parent study (**Fig. 1**). Cases were matched with controls by gestational age and sample collection dates. Gestational age remained significantly different between groups, but they were otherwise similar (**Table 1**). Three neonates were missing paired maternal samples and were excluded from maternal antibody analyses.

**Figure 1.**
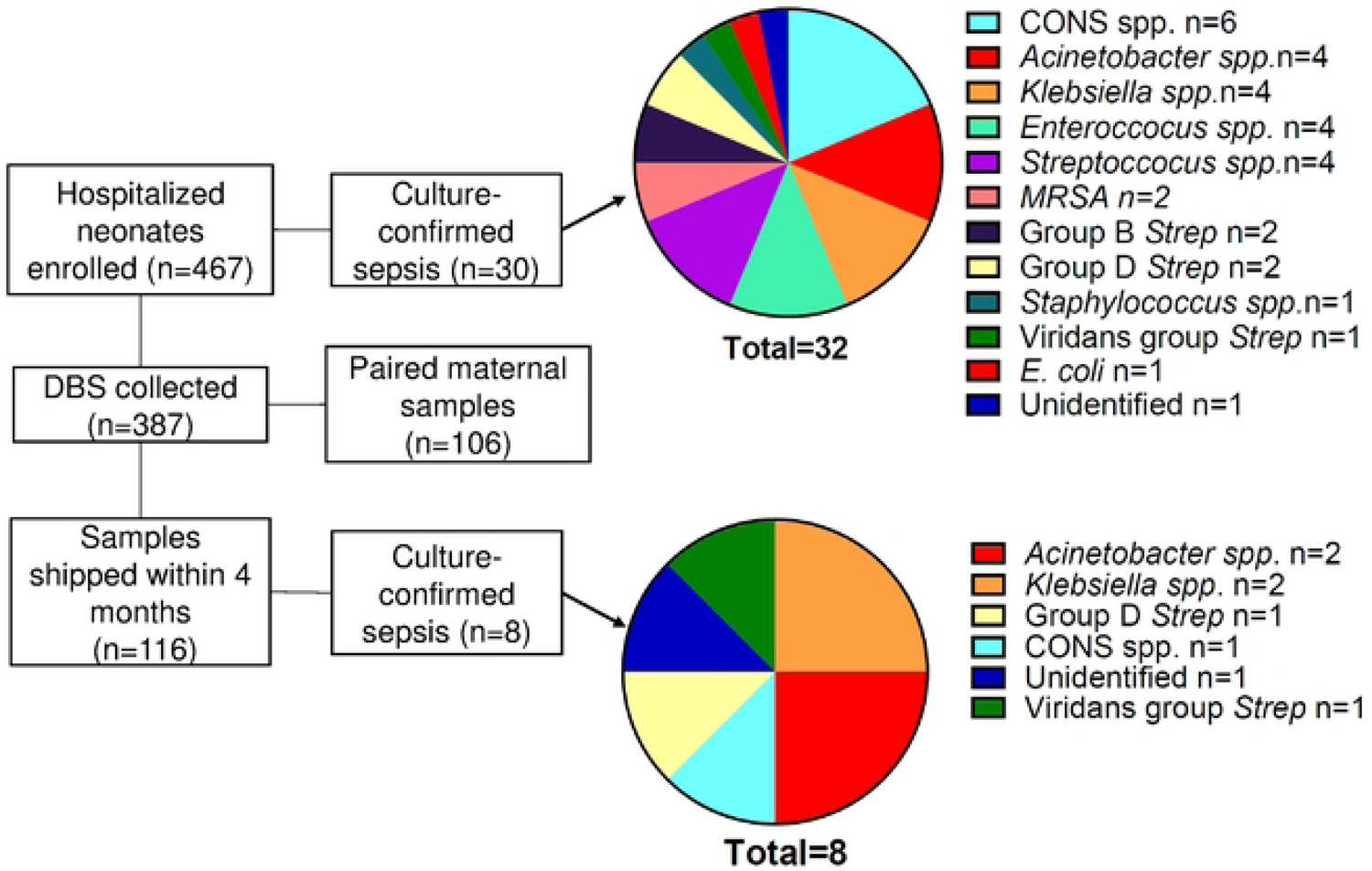
Enrollment numbers, sample numbers, and causes of sepsis in overall cohort and samples included in this analysis. CoNS= Coagulase negative *Staphyloccus spp*.; MRSA= methicillin resistant *Staphylococcus aureus*

**Table 1.**
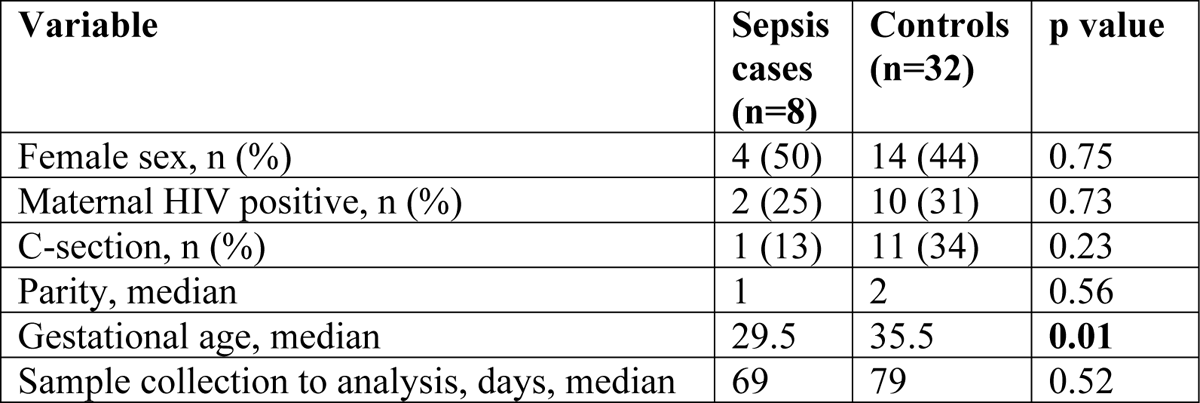
Summary of demographic variables for neonatal sepsis and control groups.

| Variable | Sepsis cases (n=8) | Controls (n=32) | p value |
| --- | --- | --- | --- |
| Female sex, n (%) | 4 (50) | 14 (44) | 0.75 |
| Maternal HIV positive, n (%) | 2 (25) | 10 (31) | 0.73 |
| C-section, n (%) | 1 (13) | 11 (34) | 0.23 |
| Parity, median | 1 | 2 | 0.56 |
| Gestational age, median | 29.5 | 35.5 | **0.01** |
| Sample collection to analysis, days, median | 69 | 79 | 0.52 |

### Establishing DBS sample integrity

Establishing whether DBS cards stored at room temperature provide valid data is important for studies in settings where immediate storage at ™20°C may be unavailable. Prior research demonstrated that most proteins on DBS cards stored at room temperature start degrading after 4 weeks but remain valid for most applications for roughly 6 months, while certain proteins can be detected after much longer periods of storage.(22–24) We, therefore, included the samples with < 4 months of storage at room temperature in this analysis. We evaluated the concentration of total IgG and *Kleb-*IgG from each sample since sample collection to estimate the degree of antibody degradation over time. We recovered lower IgG concentration over time from DBS samples (Fig. 2A). As such we normalized the amount of total IgG in each sample. Therefore, we did not observe a decrease in *Kleb-*IgG over time (Fig. 2B). Degradation of samples during room temperature storage was similar in cases compared to controls **(Fig. S1)**.

**Figure 2.**
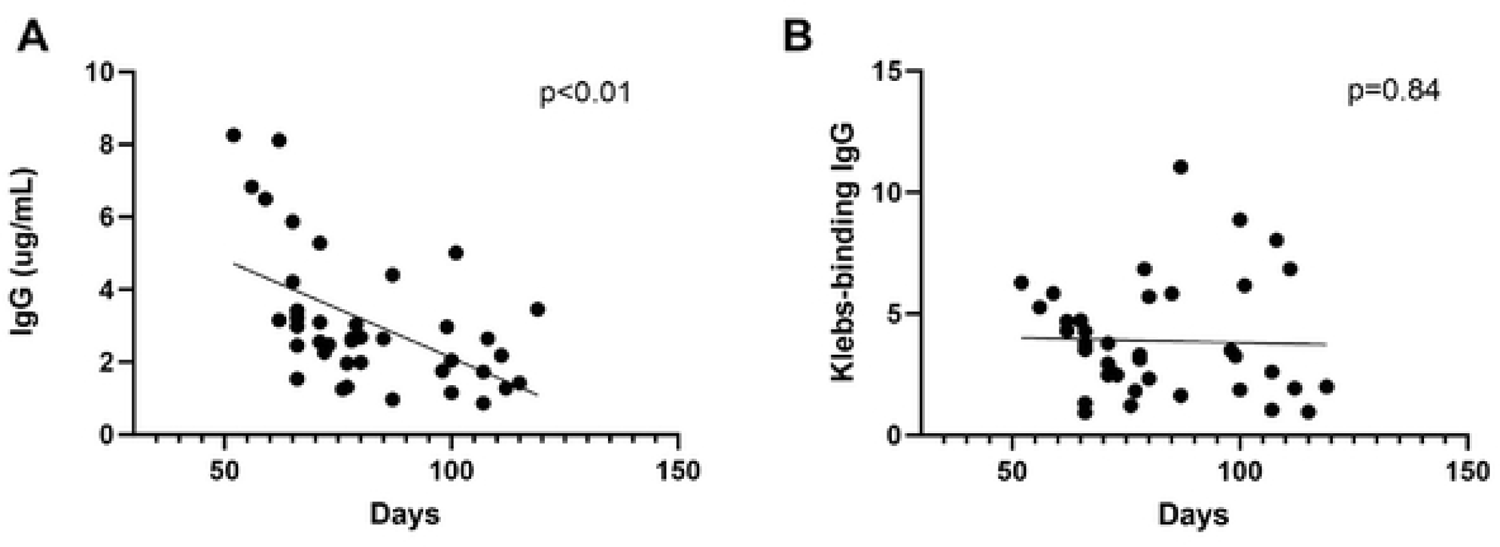
IgG concentration in DBS samples collected within 4 months. **A)** Total IgG concentration recovered from each sample as a function of time interval between sample collection and extraction. **B)** Relative amount of *Kleb-*IgG in each DBS sample, normalized to 0.4ng/µL of total IgG, as a function of time interval between sample collection and extraction. Standard Unit (SU). Simple linear regression.

As a quality check for both maternal and neonatal sample integrity, we quantified the amount of immunoglobulin A (IgA) recovered from DBS. IgA is not transplacentally transferred nor is it produced by neonates in the first weeks of life. As predicted, maternal DBS samples had detectable serum concentrations of IgA while IgA was not detected in any neonatal samples (**Fig. S2**).

### Neonates with laboratory-confirmed sepsis have lower levels of *Kleb-*IgG

We next investigated whether levels of maternal and neonatal *Kleb-*IgG were associated with increased risk for neonatal sepsis in this cohort of 8 neonates with sepsis and 32 controls without sepsis. We found significantly lower levels of *Kleb-*IgG in neonates who developed sepsis compared to neonates who did not develop sepsis (p=0.012, Fig. 3A). *Kleb-*IgG levels were lower in neonates with sepsis regardless of etiology of sepsis (p=0.04) and not significantly different in samples from neonates with sepsis due to *Klebsiella* compared to other pathogens (Fig. 3B). Since neonatal IgG concentration and specificities reflect maternal serum IgG, we compared the *Kleb-* IgG levels in samples from mothers of neonates with and without sepsis; samples from mothers of neonates who developed sepsis tended to have less *Kleb-*IgG compared to mothers of controls (p=0.06, Fig. 3C). Overall, neonatal and maternal *Kleb-*IgG levels were correlated (r=0.27, p<0.01, data not shown). In an exploratory analysis, we unexpectedly observed that there appeared to be an inverse relationship between neonatal and maternal *Kleb-*IgG in dyad samples from patients with sepsis while a positive association was apparent between neonatal and maternal *Kleb-*IgG levels in those without sepsis. **(Fig. S3).**

**Figure 3.**
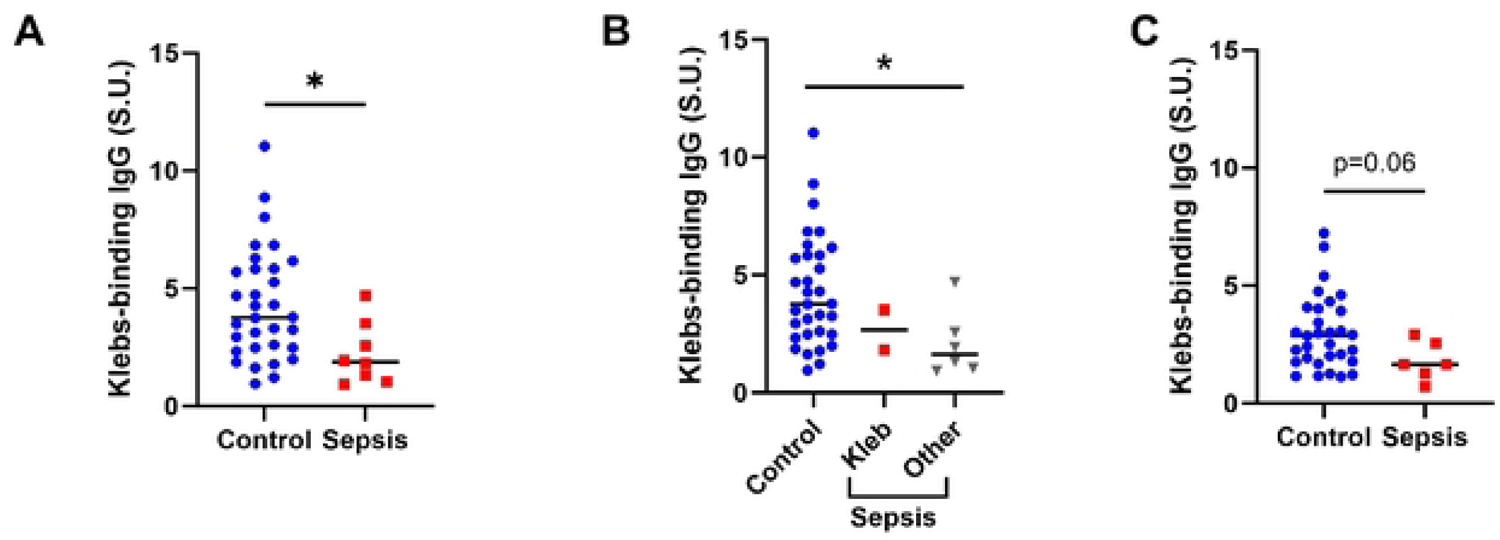
*Kleb-*IgG in neonatal and maternal samples of cases with sepsis compared to controls. **A-B)** *Kleb-*IgG binding in neonates who developed sepsis compared to matched controls. Data were normalized to standard unit, defined as the amount of *Kleb-*IgG in 100μl of reference serum (healthy adult) with 0.5 μg/mL IgG. Mann-Whitney U test (*p <.05). **C)** *Kleb-*IgG binding in serum from mothers of neonates who developed sepsis compared to mothers of controls. Kruskal Wallis test (*p<0.05).

The association of low *Kleb-*IgG levels with risk for neonatal sepsis from *Klebsiella* or other bacteria (Fig. 3B) suggests that *Kleb-*IgG is likely non-specific and may offer protection through cross-reactivity with common bacterial epitopes. Recent murine studies demonstrated that serum IgG antibodies that bind to commensal microbes protect against bacteremia from pathogenic bacteria.(25, 26) LPS is the most abundant surface molecule in most Gram-negative bacteria, and anti-LPS IgG antibodies can bind to LPS on a variety of microbes.(27) The levels of anti-LPS antibodies are associated with the risk for developing sepsis.(28) Similarly, flagellin is a virulence factor present on many bacteria and a common IgG-targeted epitope. Thus, we evaluated the degree to which IgG from each sample binds to lipopolysaccharides (LPS) and flagellin. We found similar levels of anti-LPS IgG antibodies and anti-flagellin IgG antibodies in neonates with sepsis compared to controls (**Fig. S4**). There was no association of *Kleb-*IgG levels with other potential confounders including sex, parity, maternal HIV status, or delivery mode (**Fig. S5**).

### Low *Kleb-*IgG levels are associated with neonatal sepsis across gestational ages

Given the differences in median gestational age in controls and sepsis cases, we performed a stratified analysis and found that neonates born at <33 weeks have significantly lower *Kleb-*IgG levels compared to neonates born at ≥37 weeks (p=0.03) (Fig. 4A). We evaluated this association in cases compared to controls and found no statistically significant difference in *Kleb-*IgG levels in premature neonates with sepsis compared to controls, yet a trend towards lower levels samples from neonates born at <33 weeks and between 33-37 weeks (Fig. 4B). Indeed, in an adjusted analysis, *Kleb-*IgG was associated with a decreased odds ratio (OR) of neonatal sepsis when adjusting for gestational age and days since sample collection (aOR 0.49, 95% CI [0.18, 0.93]). These analyses indicate that low Kleb-IgG levels are associated with neonatal sepsis across gestational ages.

**Figure 4.**
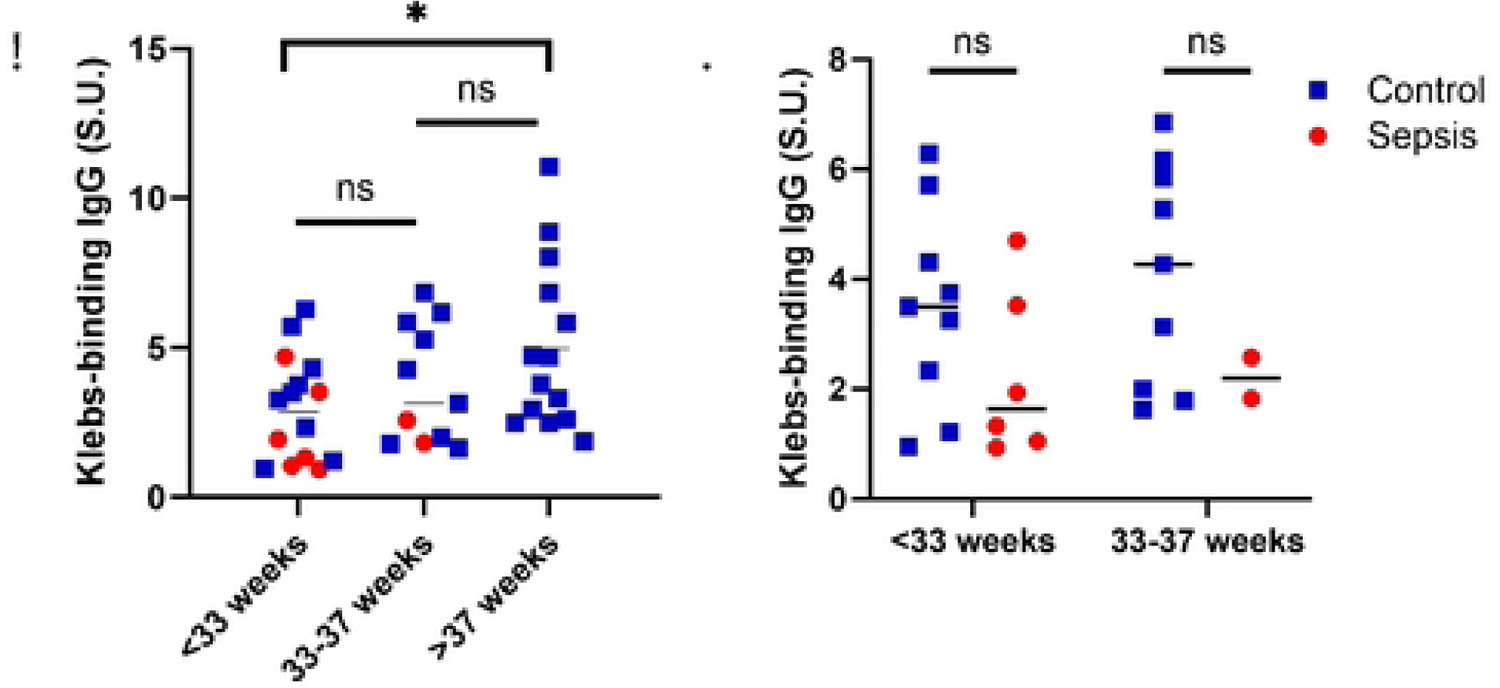
*Kleb-*IgG level by gestational age. **A)** Comparison of *Kleb-*IgG binding in neonates stratified by gestational age. Kruskal Wallis and Mann-Whitney U test (*p<0.05). **B)** Comparison of *Kleb-*IgG binding in neonates with sepsis compared to controls, stratified by gestational age. Mann-Whitney U test.

## DISCUSSION

We report an association between the level of neonatal and maternal serum IgG that bind to *K. pneumoniae* and all-cause laboratory-confirmed neonatal sepsis in a cohort of hospitalized neonates in Botswana. Using an ELISA to detect serum antibody reactivity to a *K. pneumoniae* blood culture isolate from Gaborone, Botswana, we found that neonates who developed sepsis and their mothers have lower levels of *Kleb-*IgG compared to controls when adjusting for gestational age and days of sample storage.

*K. pneumoniae* was historically the predominant isolate in blood cultures at this site, (7) leading to the development of an ELISA against a *K. pneumoniae* isolate from this NICU. During enrollment, the epidemiology of neonatal sepsis shifted from predominantly *Klebsiella* spp. to a more diverse set of microbes (Fig. 1). Remarkably, neonates with sepsis due to non-*Klebsiella* bacteria also have lower *Kleb-*IgG compared to controls suggesting that *Kleb-*IgG might provide cross-protection against common bacterial epitopes. However, neither anti-LPS nor anti-flagellin IgG antibody levels differed between neonates with sepsis compared to controls, which argues against either being an important cross-reactive microbial epitope in this cohort. These findings suggest the possibility that cross-reactivity to other common epitopes or that multiple epitopes function together to provide protection from neonatal sepsis from a range of pathogens.

By standardizing the total amount of IgG analyzed from each DBS sample, we expected to recover similar levels of *Kleb-*IgG across gestational ages, as we assumed that total IgG and pathogen-specific IgG would cross the placenta at the same rate. However, *Kleb-*IgG levels were lower in preterm neonates which may result from colonization with *Klebsiella* spp. later in pregnancy or increased production of anti-commensal antibodies later in gestation. *Klebsiella* antibodies may vary among pregnant people due to prior infections and differential abundance in the microbiome based on exposure, antibiotics, and other factors.(29) Similarly, Stach *et al* reported increased *Kleb-*IgG with increasing birthweight, a variable that is often colinear with gestational age.(30) Interestingly, another study focusing on pathogen-specific antibodies found no difference in anti-*Klebsiella* IgG by gestational age category.(31) Neither of these studies reported sepsis as an outcome.

Although there were lower levels of *Kleb-*IgG in preterm neonates in our study, the risk of sepsis remained significant when adjusting for GA and days since sample collection indicating that prematurity alone does not explain the association between lower *Kleb-*IgG levels and neonatal sepsis. Since the rate of sample degradation was similar between cases and controls, this is unlikely to account for the association (**Fig. S1**). In an exploratory analysis, we noted a positive linear correlation of maternal and neonatal *Kleb-*IgG among the whole cohort regardless of gestational age, which has been reported previously.(31) However, when we separated cases and controls, we noted an inverse correlation in dyads with sepsis and a positive correlation in control dyads which may indicate different rates of placental transfer (**Fig. S3**). Many factors are known to influence transplacental antibody transfer including parity, history of infections, timing of vaccination and placental insufficiency.(14, 32) Data on placental health, prior infections and maternal hypertension were not available in this cohort, thus we are unable to compare with these findings.

Our study highlights the benefits of using DBS for sample collection. Compared to traditional blood tests, DBS can be obtained via minimally invasive methods (heel stick vs. venipuncture), requires a minimal blood volume, remains stable when stored at room temperature for up to 4 months, can be transported easily with minimal risk for contamination, and is an efficient and feasible method to acquire serum samples for a variety of assays.(33, 34) However, there are limitations including variation in collection volume and sample degradation which unfortunately limited the sample size in this study.

## CONCLUSIONS

We propose that maternal IgG antibodies against *K. pneumoniae* are transferred *in utero* late in gestation and may be associated with a decreased risk of sepsis from diverse pathogens. These findings suggest species-specific maternal antibodies may confer broader protection against common neonatal pathogens. This supports the ongoing development of vaccination strategies during pregnancy to increase pathogen-binding IgG levels in neonates. Further studies are needed to explore mechanisms and enhance efficient transfer of protective antibodies to premature neonates.

## Data Availability

The data used in the reported analyses is available as a supplementary document with this article.

## Acknowledgements

The authors thank Mrs. Relebogile Thipe for helpful input with IRBs. The authors also thank the study participants and their families, as well as the staff in the Neonatal Unit. This publication was made possible through core services and support from the Penn Center for AIDS Research (CFAR), an NIH-funded program (P30 AI 045008).

## Contributorship Statement

S.Z. and C.M.M. contributed equally to this paper. Conceptualization, S.E.C., C.M.M., S.Z. and M.A.S.; Methodology, S.Z. and T. D.; Writing – Original Draft, S.Z. and M.A.S.; Writing – Review & Editing, S.Z., T.D., C.M.M., J.S., A.P.S., M.R.G., S.E.C. and M.A.S..; Conceptualization of parent study, C.M.M., J.S., O.B., T.A-M., A.P.S., S.E.C.; Enrollment of patients and data collection, J.S., B.M., N.T.; Provided care for study patients, J.S., B.N., O.B., A.G.; Data review, S.Z., T.D., C.M.M., J.S., A.P.S., B.N., T.A-M., M.R-G., P.J.P., S.E.C., M.A.S; Funding Acquisition, S.E.C., C.M.M., and M.A.S.; Resources, M.A.S.; Supervision, M.A.S. All authors reviewed the final version of the manuscript prior to submission.

---

**Supplementary Figure 1.**
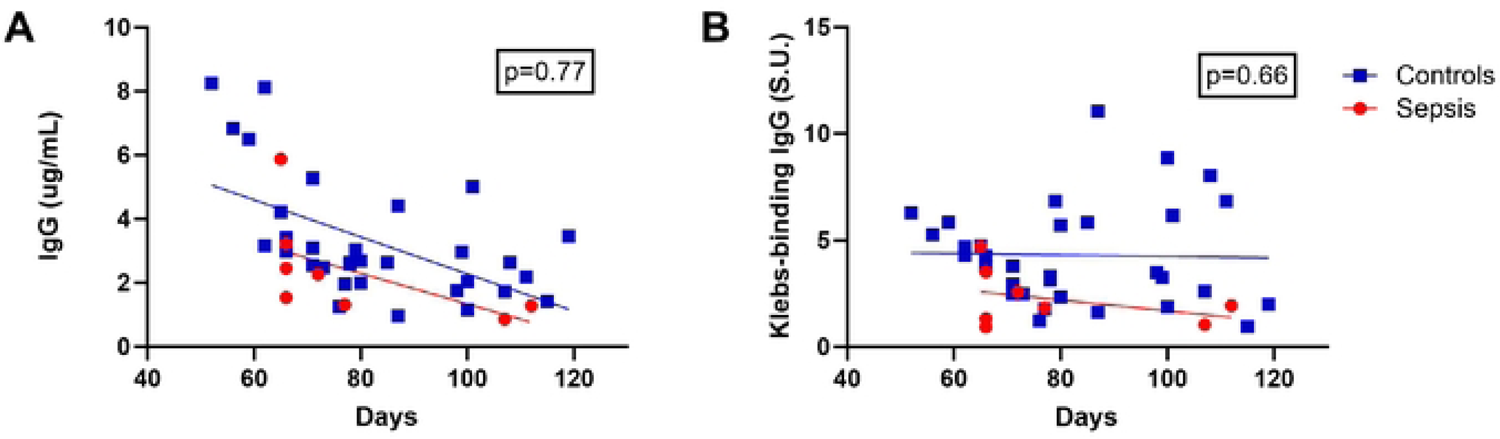
Immunoglobulin levels over time in cases compared to controls. **A)** Total IgG concentration recovered from each sample as a function of time interval between sample collection and extraction in sepsis cases compared to controls. **B)** Relative amount of *Kleb-*IgG in each DBS sample as a function of time interval between sample collection and extraction. Standard Unit (SU). Slopes were compared by analysis of covariance.

**Supplementary Figure 2.**
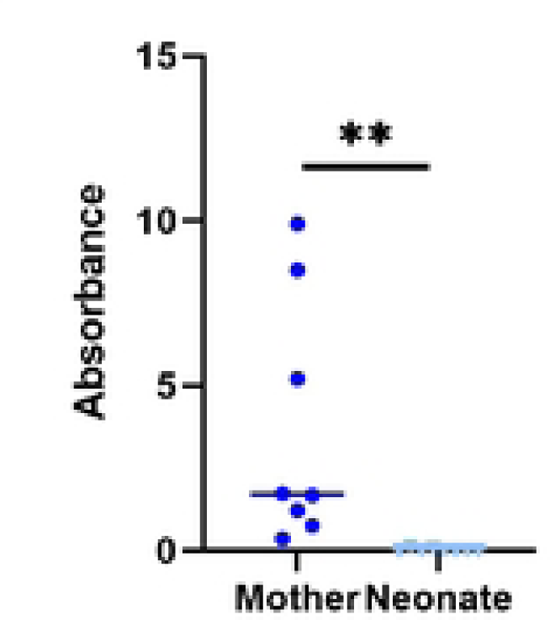
Levels of IgA in maternal and neonatal DBS samples. IgA level was determined using standard human ELISA from randomly selected paired maternal and neonatal samples. Mann-Whitney U test. **p<0.01.

**Supplementary Figure 3.**
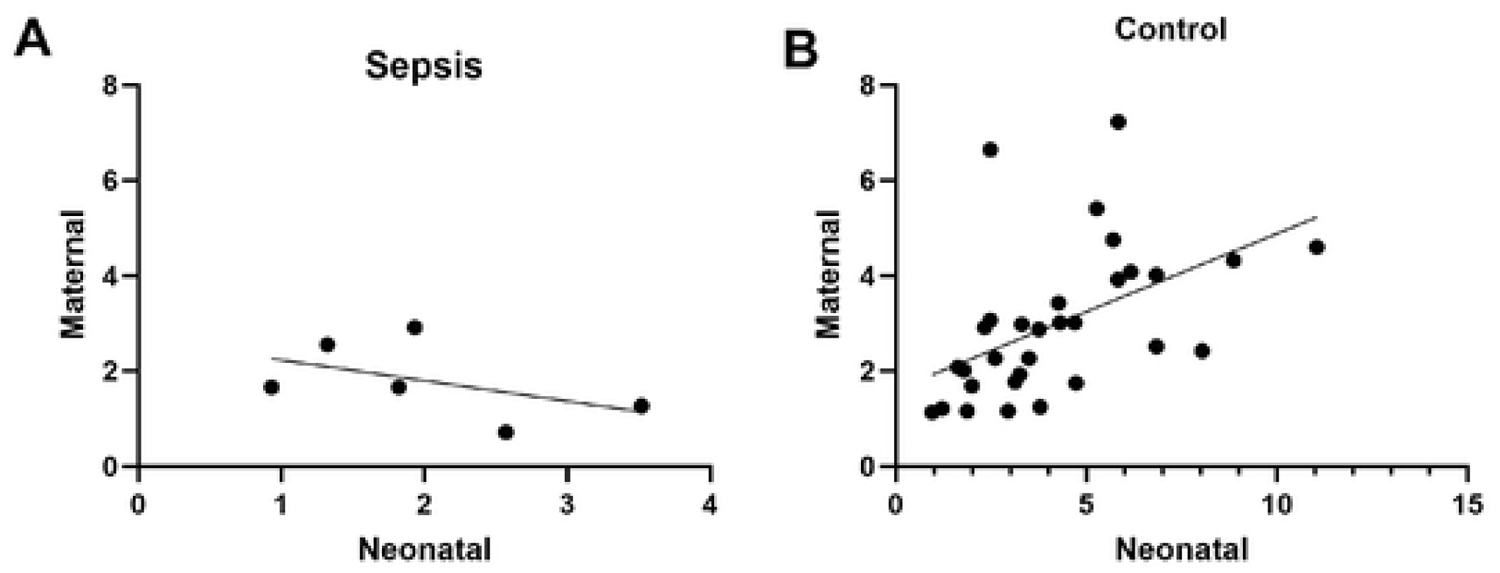
Correlation between maternal *Kleb-*IgG and neonatal *Kleb-*IgG levels. **A)** *Kleb-*IgG levels for mother-neonate dyads with sepsis, simple linear regression, p=0.32. **B)** *Kleb-*IgG levels for mother-neonate dyads with sepsis, simple linear regression, p<0.01.

**Supplementary Figure 4.**
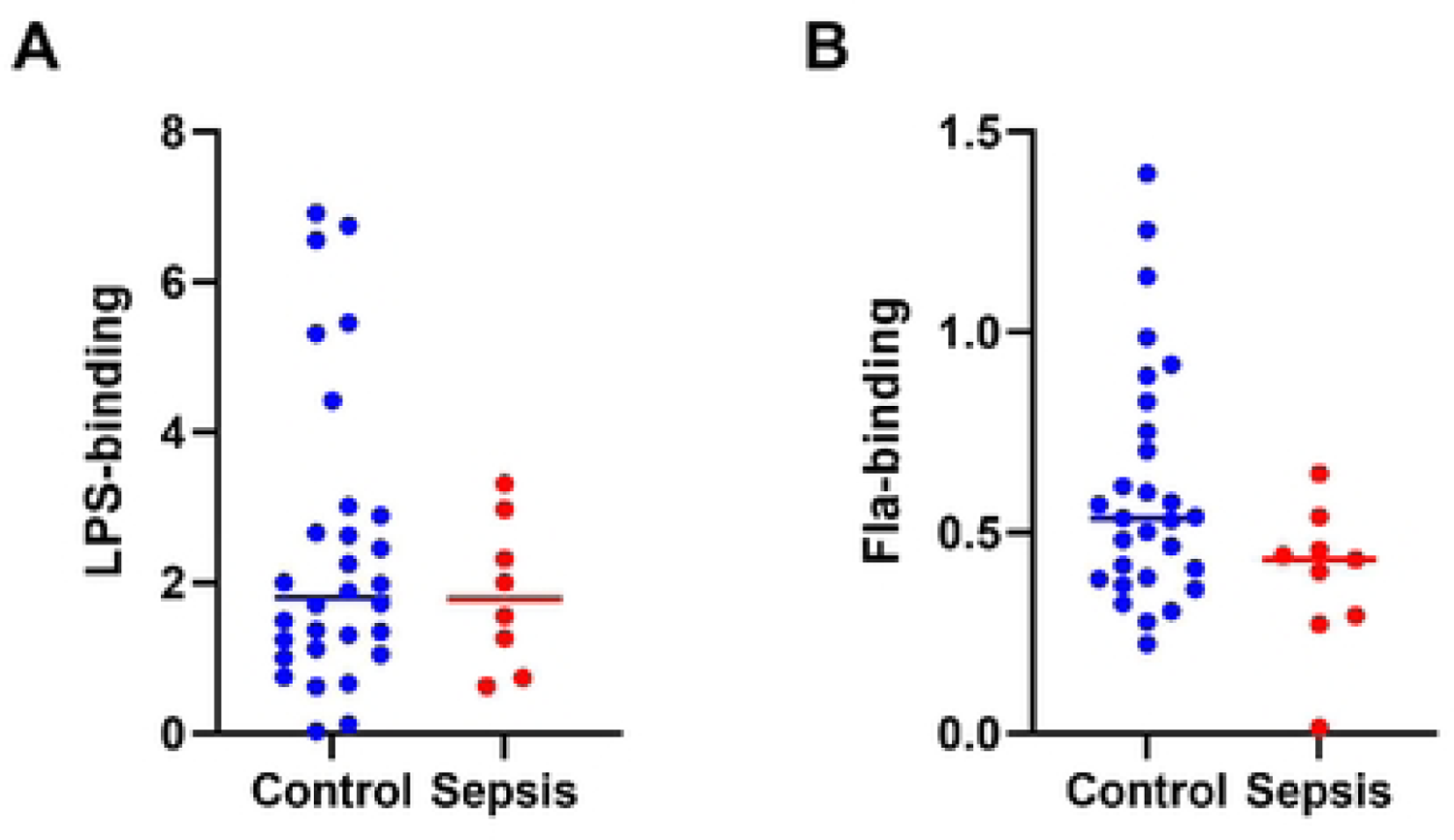
Anti-LPS IgG and anti-flagellin IgG in neonates with and without sepsis. **A)** ELISA comparing anti-LPS IgG in neonates with sepsis vs. controls. All units are normalized to the standard unit, defined as the amount of anti-LPS IgG in 50 ng of reference adult serum. **B)** ELISA comparing anti-flagellin IgG in neonates with sepsis vs. controls. Mann-Whitney U test.

**Supplementary Figure 5.**
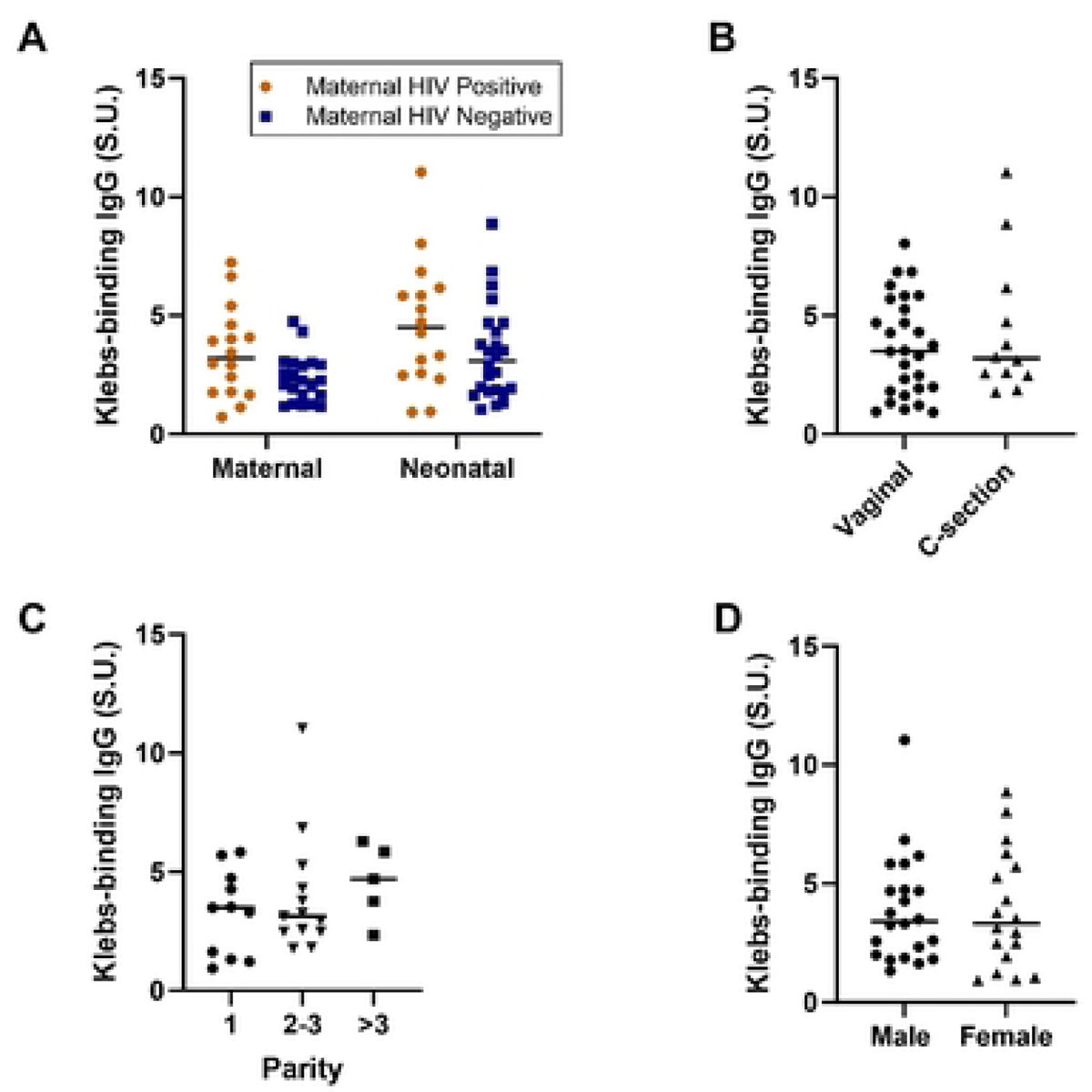
*Kleb-*IgG level by maternal HIV status, neonatal sex, delivery mode, and maternal parity. A-D) *Kleb-*IgG binding comparison by the indicated clinical variables. Mann-Whitney U test or Kruskal-Wallis test.

